# Modulations in high-density EEG during the suppression of phantom-limb pain with neurostimulation in upper-limb amputees

**DOI:** 10.1101/2023.08.13.23294037

**Authors:** Daria Kleeva, Gurgen Soghoyan, Artur Biktimirov, Nikita Piliugin, Yury Matvienko, Mikhail Sintsov, Mikhail Lebedev

## Abstract

Phantom limb pain (PLP) is a distressing and persistent sensation that occurs after the amputation of a limb. While medication-based treatments have limitations and adverse effects, neurostimulation is a promising alternative approach whose mechanism of action needs research, including electroencephalographic (EEG) recordings for the assessment of cortical manifestation of PLP relieving effects. Here we collected and analyzed high-density EEG data in three patients (P01, P02, and P03). Peripheral nerve stimulation (PNS) suppressed PLP in P01 but was ineffective in P02. By contrast, transcutaneous electrical nerve stimulation (TENS) was effective in P02. In P03, spinal cord stimulation (SCS) was used to suppress PLP. Changes in EEG oscillatory components were analyzed using spectral analysis and Petrosian fractal dimension (FD). With these methods, changes in EEG spatio-spectral components were found in the theta, alpha, and beta bands in all patients, with these effects being specific to each individual. The changes in the EEG patterns were found for both the periods when PLP level was stationary and the periods when PLP was gradually changing after neurostimulation was turned on or off. Overall, our findings align with the proposed roles of brain rhythms in thalamocortical dysrhythmia or disrubtion of excitation and inhibition which has been linked to neuropathic pain. The individual differences in the observed effects could be related to the specifics of each patient’s treatment and the unique spectral characteristics in each of them. These findings pave the way to the closed-loop systems for PLP management where neurostimulation parameters are adjusted based on EEG-derived markers.

## Introduction

Phantom limb pain (PLP) is a distressing and persistent sensation experienced after the amputation of a limb. The phenomenology of PLP is diverse, with common sensations including pressure, aching, burning, and spasms (McCormick et al., 2014). This condition has a significant impact on the quality of life of affected individuals, with the estimates of its prevalence reaching as high as 64 % (Limakatso et al., 2020), which underscores the need for effective management strategies.

Pharmacological treatment is the primary approach to managing PLP, with a range of medications available including beta-blockers, anaesthetics, opioids, NMDA receptor antagonists, muscle relaxants, nerve blocks, and others. However, the short- and long-term effectiveness of most of these medications is not well established. Even medications like morphine, gabapentin, or ketamine, which are associated with short-term pain relief, could have adverse side effects such as constipation, sedation, and respiratory problems (Alviar et al., 2016).

In light of the limitations of pharmacological interventions, neurostimulation is emerging as a promising treatment option to treat PLP, including spinal cord stimulation (SCS) and peripheral nerve stimulation (PNS) (Soghoyan et al., 2023). Both SCS and PNS activate afferents fibers while PNS is more selective to influence residual limb nerves (Petersen et al., 2019). Whereas the pain-relieving mechanisms of neurostimulation are not completely understood, several explanations have been proposed. According to the gate control theory (Melzack and Wall, 1965), the electrical stimulation of non-nociceptive large-diameter *Aβ* fibers suppresses the pain signals transmitted to the spinal cord through the smaller *Aδ* and *C* fibers by inhibiting interneurons involved in this transmission. In addition to the effects that the neurostimulation has on the spinal cord circuitry, higher hierarchical levels could be also engaged where neurostimulation-induced inputs reach cortical sensorimotor areas and offset the amputation-related remapping and pathological activity.

While the patients’ subjective reports of the changes in PLP are used as the key criterion of treatment effectiveness, neurophysiological indicators could improve the assessment of treatment effects. Thus, using electroencephalography (EEG) as an objective marker of response to neurostimulation could significantly aid the treatment and lead to the development of closed-loop systems where neurostimulation parameters are adjusted based on EEG measurements. Better understanding of how EEG patterns correspond to different stages of PLP will facilitate the development of improved diagnostics, prognostics, and treatment monitoring for PLP, as well as optimizing current neurostimulation approaches.

EEG patterns that could serve as biomarkers of pain are not sufficiently understood. According to a recent review of EEG spectral characteristics in chronic neuropathic pain (Mussigmann et al., 2022), theta power grows consistently when pain increases. Studies on the involvement of alpha and beta rhythms have yielded mixed results, with some studies showing a correlation between the power in these bands and pain intensity and others reporting an opposite trend. These differences could be related to heterogeneity in the study designs: whether the eyes were open or closed, the specific frequency bands used in the spectral analysis, and other settings.

Several studies examined EEG patterns in amputees. In a case report (Walsh et al., 2015), the left frontal spectral power increased in a patient who attempted movements of the phantom limb, and these EEG modulations resembled the ones observed in healthy subjects during voluntary movements of their limbs. A study of the resting-state EEG network (Lyu et al., 2016) compared healthy controls to right-hand amputees. Alterations were found after limb amputations in the global and local phase synchronization of the alpha and beta rhythms. In a study of non-painful electrical stimulation of the limbs in amputees, stimulation of the affected limb resulted in an increase in the response of the N/P135 dipole (Vase et al., 2012). Additionally, alpha wave coherence increased with PLP reduction during virtual reality (VR) rehabilitation and vibrotactile stimulation of the referred sensation in the cheek and shoulder (Osumi et al., 2020). Using intracranial EEG recordings from the anterior cingulate cortex and orbitofrontal cortex, Shirvalkar et al. (2023) predicted the severity of chronic pain, including PLP. They found that while certain predictors showed some consistency (such as chronic pain being associated with orbitofrontal activity and acute pain with anterior cingulate cortex activity), the spectral characteristics and the weights of these features varied across patients, which hindered generalization to a wide population of patients.

Overall, the current knowledge of EEG correlates of PLP and other types of pain is insufficient for the development of efficient bidirectional systems for pain treatment. As a step toward the knowledge in this field, this study aimed to assess EEG patterns resulting from the treatment of PLP with PNS and SCS in three patients with upper-limb amputations.

## Materials and methods

### Participants

EEG data were collected in three upper-limb amputees: patients P01, P02, and P03. All three patients signed an informed consent. The experimental design was approved by the Ethical Committee of FEFU Biomedicine school (Protocol num. 4; April 16, 2021). The study was registered as a clinical trial num. NCT05650931 on the platform ClinicalTrials.gov. P01 and P03 had transhumeral amputations on the left side, and P02 had a transradial amputation of the right side. All patients experienced PLP at the level of 7-8 points estimated by a visual analogue scale (VAS).

### Treatment

All patients were treated with neurostimulation, with individual differences in the stimulation sites and treatment strategies.

PNS was performed with 8-contact electrodes (Directional Lead for the St. Jude Medical Infinity™ DBS System; Abbot; USA) implanted in the median nerve. SCS was performed with the cylindrical electrodes (Vectris Surescan Trail MRI 1×8 compact 977D260; Medtronic; USA) implanted epidurally over the spinal cord at the Th6-7 level. The patients experienced sensations in the phantom hand during PNS or SCS. Additionally, in patient P02, TENS was performed where the electrodes were placed on the skin of the residual limb in close proximity to the ulnar nerve.

PNS of the median nerve was effective to suppress PLP in P01, so this patient was treated with an ongoing PNS. PNS of the median nerve was ineffective in P02 but transcutaneous electrical nerve stimulation (TENS) was effective. In P03, spinal cord stimulation (SCS) was applied through the electrodes implanted over the segments Th6-7 to suppress PLP.

For patient P01, PNS was applied at a frequency of 40Hz, utilizing burst mode with a frequency of 500 Hz, pulse width of 1000 microseconds, and amplitude of 0.15mA. In the case of patient P03, SCS was administered at a frequency of 100Hz, with a pulse width of 100 microseconds and an amplitude of 2.4mA. For patient P02, two stimulation modes were implemented. PNS was applied at a frequency of 50Hz, pulse width of 100 microseconds, and an amplitude of 0.9 mA. Additionally, TENS was delivered at a frequency of 100Hz, pulse width of 100 microseconds, and an amplitude of 18mA.

The assessment of pain intensity was conducted using the Visual Analogue Scale (VAS), a standard tool for quantifying subjective pain experiences. The VAS is typically represented by a straight horizontal line, usually 100 mm long. The ends of this line signify the extreme boundaries of the evaluated parameter, which could be a symptom, pain level, or a health-related aspect. The scale is oriented from left to right, with the left end representing the worst possible state and the right end indicating the best. Participants indicate their personal experience on this line, providing a measure of the intensity of the parameter in question. Additionally, the patients provided comments on the other sensations they experienced and their location on the phantom limb.

### EEG recordings

EEG signals were sampled at 1,000 Hz with an NVX-136 amplifier. A 128-channel cap was used in patients P01 and P03. In patient P02, a smaller 60-channel cap was used because it fit the head better. The ground electrode was set to AFz. Ear clips were used as reference electrodes (positions A1 and A2) at the stage of data acquisition. Subsequently, the data were re-referenced to common average reference.

In all patients, resting-state EEGs were recorded with eyes open and closed during different stages of neurostimulation procedures, including stimulation being turned on or off and the associated states of PLP. The additional set of the recordings were intended to capture gradual changes in EEG patterns after the stimulation was turned on or off. For each condition, the participants verbally described their sensations. The audio of these comments was recorded. For the nonstationary conditions where PLP was gradually changing after the stimulation was turned on or off, EEG recordings continued until the PLP level stabilized.

In patient P01, PNS continued throughout the day and was powerful enough to remove PLP completely. Accordingly, EEG recordings started with the stationary condition where PNS was still ongoing and no PLP was present. Next, after the eye-open and eye-closed data were collected, PNS was turned off. The subsequent EEG recordings corresponded to a nonstationary condition where PLP was gradually reappearing. After the patient reported that PLP stabilized, resting-state EEG was recorded for the PLP experienced in the absence of PNS. After this recording was completed, PNS was turned on, after which PLP was gradually disappearing.

In patient P02, EEG data were collected for both PNS, ineffective for PLP relief, and TENS which was effective. The PNS dataset contained data for three conditions: PNS on, PNS off, and PNS on again. The TENS dataset covered the following conditions: TENS off, TENS on, and TENS off again. Periods were documented where PLP was gradually decreasing after TENS was turned on and increasing after it was turned off.

Patient P03, in whom SCS was used to suppress PLP, was available only for one recording session. The patient took a pain-relieving medication on the night prior to the session, so no PLP was experienced in the beginning of the recordings although SCS was turned off. Next, while though there was still no PLP because of the medication effect, SCS was turned on. Finally, SCS was turned off, but this time the patient started to experience PLP.

### EEG analysis

EEG was analyzed in MNE Python (Gramfort et al., 2013). The recordings were band-pass filtered between 3 and 30 Hz. Noisy and flat channels were discarded manually and interpolated where appropriate. The EOG, muscle, powerline, and neurostimulation artifacts were removed by the means of independent component analysis (ICA, FastICA algorithm). Next, we conducted a spectral analysis of the cleaned signals, which consisted of calculating the relative power spectral density using multitaper on 10-s segments of the EEG data.

To compare the properties of oscillatory components of interest, we conducted spatio-spectral decomposition (SSD) for periodic components of the spectra (Nikulin et al., 2011). This method involves a linear decomposition of multichannel EEG data which optimizes the signal-to-noise ratio by maximizing the peak-frequency power and minimizing it for the neighboring bands. For our study, we defined these neighboring bands as a range of ±4 Hz around the targeted frequency band.

Because of the variability in the EEG spectral profiles across the participants, we assessed the EEG patterns with the appropriate for such cases metric called Petrosian fractal dimension (FD) (Petrosian, 1995) , which was computed on 30-s EEG segments. The Petrosian FD was computed as:

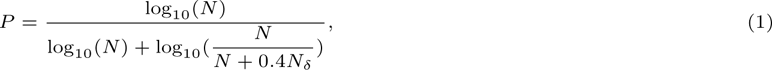

where *N* is the length of a time series for a single EEG channel, and *N_δ_* denotes the count of sign changes in the derivative of the signal. This approach was developed for the estimation of EEG signal complexity. The FD measure matches well the other complexity metrics such as different types of entropy and Lempel-Ziv complexity.

### Statistical analysis

Since we analyzed each patient individually and the spectral properties of resting-state EEG were variable across the conditions, we did not explicitly divide power spectral density (PSD) into frequency bands. In order to determine PSD segments and channels with statistically significant changes, we used a permutation cluster test Maris and Oostenveld (2007), which resolved the multiple comparisons problem for the features in question (a set of frequencies and channels). In this test, the cluster-forming threshold was set to 0.01*/*2 for the observed t-values because the statistics was two-tailed. This level of significance was chosen to strike a balance between localized and generalized effects. When there were multiple comparisons, such as in cases where we compared conditions with PLP present, PLP absent due to stimulation, or PLP present due to ineffective stimulation, we adjusted this threshold to account for the number of comparisons. The number of permutations was set to 10^3^. In instances involving multiple comparisons, the cluster p-values were adjusted accordingly using Bonferroni correction.

To maintain the continuity criterion, clusters that comprised less than 10 % of the total number of channels and spanned a range of less than 1.5 Hz were discarded.

For the FD data, a similar approach was employed, setting the cluster-forming threshold at 0.05*/*2 and additionally demeaning the FD values within each epoch of the dataset.

To analyze EEG patterns for the cases where PLP changed gradually, we used a Mann-Kendall test, which detected the features characterized by significant monotonic trends, and reported the PSD values for each time instance averaged over the channels where the trends were statistically significant.

## Results

### Subjective reports

The subjective experiences of each of the patient were highly individual.

In patient P01, PNS strongly suppressed PLP with some day to day variations of this effect. When EEG recordings were conducted in this patient, 90 % PLP suppression was reported. In the absence of PNS, PLP was felt in the phantom of the left wrist and was accompanied by a sensation of compression. Both of these feelings were suppressed by PNS. PNS evoked sensations itself in the phantom that were localized to the left palm, in the area of the dermatome C6.

In patient P02, PNS did not suppress PLP, but TENS did. On the recording day when PNS was tested, PLP was felt in the right little finger and right wrist. PNS induced phantom sensations in the forearm in the area normally innervated by the right lateral antebrachial cutaneous nerve. On the day when TENS was tested and was effective to suppress PLP, the patient reported PLP suppression by 60 % in the right little finger and the right bend of the elbow. TENS evoked phantom sensations at the base of the right thumb.

In patient P03, PLP was felt over the entire left palm. SCS evoked sensations in the same area of the phantom and suppressed PLP by 10-30 %. On the day when EEG recordings were conducted, the patient took medication which abolished PLP completely, but during the last recording session when the SCS was turned off, PLP returned to the usual level.

### Spatio-spectral characteristics

The analysis of the EEG spatio-spectral characteristics revealed across-condition modulations. Figure 1 and Table 1 show the results of the PSD analysis for the eyes-open and eyes-closed conditions.

**Fig. 1.**
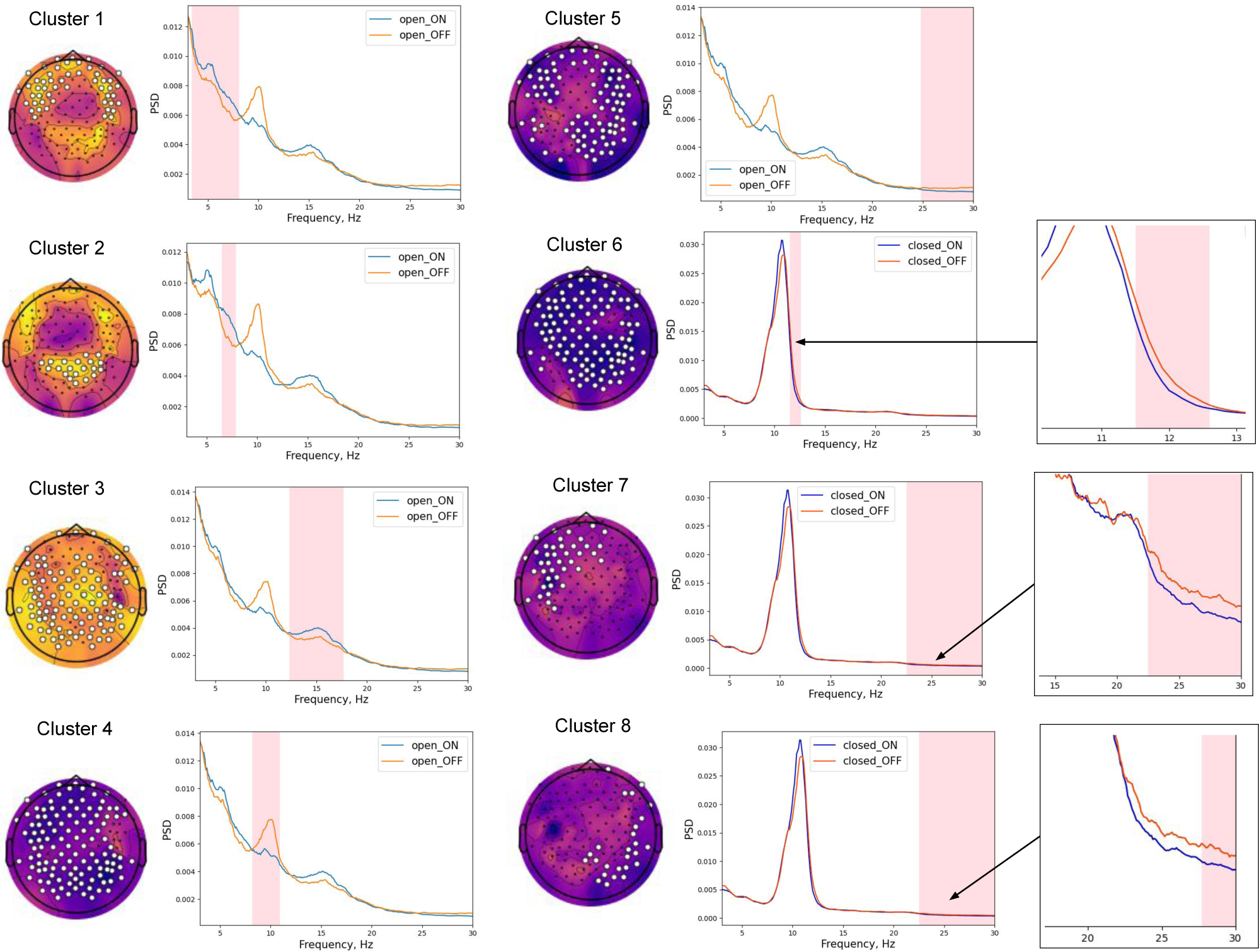
Spatio-spectral clusters in patient P01 with significant changes after PNS was turned off and PLP returned. Frequency intervals with significant changes in spectral power are shaded in pink; EEG channels, forming the clusters with significant differences, are marked with white dots. Scalp topographies depict the t-values

**Table 1.**
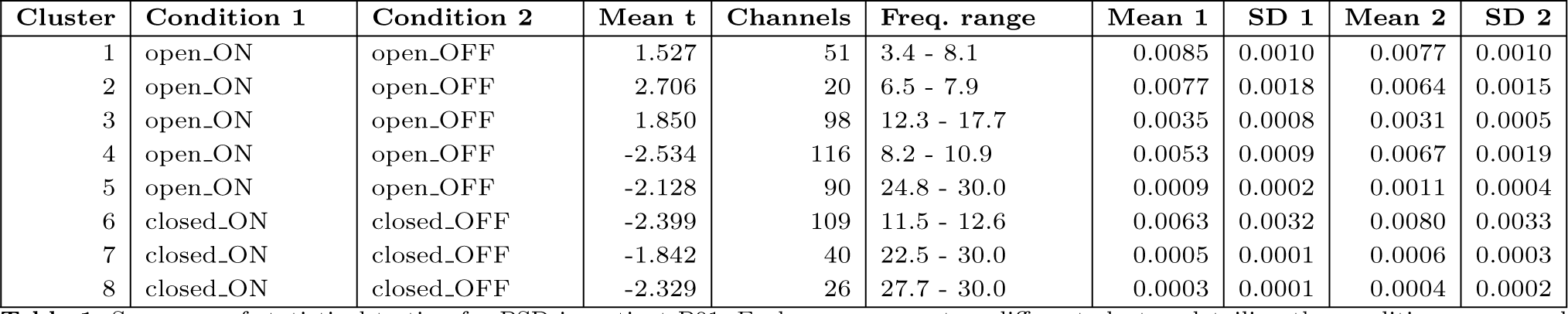
Summary of statistical testing for PSD in patient P01. Each row represents a different cluster, detailing the conditions compared (Condition 1 and Condition 2), mean t-values, number of channels involved, frequency range of the cluster, and the mean with standard deviation for both conditions

| Cluster | Condition 1 | Condition 2 | Mean t | Channels | Freq. range | Mean 1 | SD 1 | Mean 2 | SD 2 |
| --- | --- | --- | --- | --- | --- | --- | --- | --- | --- |
| 1 | open_ON | open_OFF | 1.527 | 51 | 3.4 - 8.1 | 0.0085 | 0.0010 | 0.0077 | 0.0010 |
| 2 | open_ON | open_OFF | 2.706 | 20 | 6.5 - 7.9 | 0.0077 | 0.0018 | 0.0064 | 0.0015 |
| 3 | open_ON | open_OFF | 1.850 | 98 | 12.3 - 17.7 | 0.0035 | 0.0008 | 0.0031 | 0.0005 |
| 4 | open_ON | open_OFF | -2.534 | 116 | 8.2 - 10.9 | 0.0053 | 0.0009 | 0.0067 | 0.0019 |
| 5 | open_ON | open_OFF | -2.128 | 90 | 24.8 - 30.0 | 0.0009 | 0.0002 | 0.0011 | 0.0004 |
| 6 | closed_ON | closed_OFF | -2.399 | 109 | 11.5 - 12.6 | 0.0063 | 0.0032 | 0.0080 | 0.0033 |
| 7 | closed_ON | closed_OFF | -1.842 | 40 | 22.5 - 30.0 | 0.0005 | 0.0001 | 0.0006 | 0.0003 |
| 8 | closed_ON | closed_OFF | -2.329 | 26 | 27.7 - 30.0 | 0.0003 | 0.0001 | 0.0004 | 0.0002 |

**Table 2.** Summary of statistical testing for PSD in patient P02 (eyes-open conditions). Each row represents a different cluster, detailing the conditions compared (Condition 1 and Condition 2), mean t-values, number of channels involved, frequency range of the cluster, and the mean with standard deviation for both conditions. ‘open ON‘ condition represents TENS turned on, while ‘open ON (ineff.)‘ represents PNS, which was ineffective for PLP suppression, turned on

| Cluster | Condition 1 | Condition 2 | Mean t | Channels | Freq. range | Mean 1 | SD 1 | Mean 2 | SD 2 |
| --- | --- | --- | --- | --- | --- | --- | --- | --- | --- |
| 1 | open_ON | open_OFF | 2.428 | 9 | 20.7 - 24.7 | 0.0031 | 0.0006 | 0.0026 | 0.0005 |
| 2 | open_ON | open_OFF | 3.153 | 53 | 25.1 - 30.0 | 0.0016 | 0.0003 | 0.0013 | 0.0002 |
| 3 | open_ON | open_OFF | -1.986 | 46 | 4.6 - 8.6 | 0.0031 | 0.0005 | 0.0036 | 0.0005 |
| 4 | open_ON (ineff.) | open_OFF | 1.165 | 7 | 3.0 - 9.1 | 0.0032 | 0.0005 | 0.0030 | 0.0004 |
| 5 | open_ON (ineff.) | open_OFF | 2.170 | 48 | 19.4 - 30.0 | 0.0025 | 0.0003 | 0.0022 | 0.0002 |
| 6 | open_ON (ineff.) | open_OFF | -1.695 | 42 | 3.0 - 10.2 | 0.0032 | 0.0003 | 0.0035 | 0.0004 |
| 7 | open_ON (ineff.) | open_OFF | -1.516 | 14 | 9.7 - 17.8 | 0.0056 | 0.0006 | 0.0061 | 0.0006 |
| 8 | open_ON (ineff.) | open_OFF | -2.325 | 14 | 15.3 - 18.2 | 0.0057 | 0.0008 | 0.0064 | 0.0007 |
| 9 | open_ON | open_ON (ineff.) | 1.313 | 12 | 3.2 - 8.3 | 0.0033 | 0.0006 | 0.0031 | 0.0004 |
| 10 | open_ON | open_ON (ineff.) | 2.110 | 7 | 12.7 - 16.8 | 0.0072 | 0.0008 | 0.0064 | 0.0008 |
| 11 | open_ON | open_ON (ineff.) | 2.059 | 28 | 25.1 - 30.0 | 0.0017 | 0.0003 | 0.0015 | 0.0003 |
| 12 | open_ON | open_ON (ineff.) | 3.154 | 12 | 28.5 - 30.0 | 0.0013 | 0.0004 | 0.0011 | 0.0003 |
| 13 | open_ON | open_ON (ineff.) | -1.895 | 10 | 3.0 - 9.1 | 0.0025 | 0.0003 | 0.0029 | 0.0004 |

One noticeable change for the eyes-open condition was the presence of a prominent alpha peak as PNS was turned off and PLP was present. The analysis attributed this effect to cluster 4. The spatial distribution of t-values clearly indicates that the observed difference predominantly manifested in the anterior channels (ipsilateral to the amputated side) and posterior channels (contralateral to the amputated side). However, the cluster encompassed nearly all channels, demonstrating that the effect is widespread and generalized.

When the PNS was turned on, increases occurred in the theta (clusters 1 and 2) and low beta power (cluster 3, 12.3-17.7 Hz). Specifically, cluster 1 was localized in the frontal channels, cluster 2 was predominant over the parietal channels, and cluster 3 demonstrated a spatially widespread pattern.

Additionally, we observed pain-related increase in high-beta power (cluster 5, 24.8-30 Hz), when PNS was turned off. The cluster involved the frontal regions, and additionally, it extended to the temporal and posterior areas, predominantly contralateral to the side of amputation.

When patient P01 closed the eyes, the alpha rhythm prominently increased. Three clusters, clusters 7, 8, and 9, demonstrated significant changes as PNS was turned off. For cluster 7 a generalized increase in the higher range of the alpha band (11.5-12.6 Hz) was found. For clusters 8 and 9 an increase in the high beta power (22.5-30 Hz) was found in the frontal areas ipsilateral to the amputation and posterior areas contralateral to the amputation, which resembled the changes for cluster 5 in the eyes open condition.

We observed gradual changes in EEG patterns in patient P01 occurring after PNS was turned off or on. In general, these changes matched the gradual changes in PLP that the patient reported. To quantify the gradual changes in EEG, we examined the mean PSD for the presence of linear trends within the frequency bands identified in the analysis of spatio-spectral clusters. First, we analyzed the time course of alpha power, which was elevated in the absence of PNS. We identified a significant increasing trend for 92 channels (mean slope: 8.497e-06, mean intercept: 0.0047) (Figure 2, 1.A). These channels matched the channels with the alpha-rhythm increase identified in the analyses of epochs with the stationary level of PLP (cluster 4 in Figure 1). Notably, a sharp increase occurred in the alpha rhythm toward the end of the nonstationary period. Yet, even when the period of this change was excluded the trend remained evident, (Figure 2, 1.B). For this shorter analysis epoch, 17 channels had a significant increasing trend (mean slope: 8.198e-06, mean intercept: 0.0047), and the majority of these channels were located in the anterior cortical regions.

**Fig. 2.**
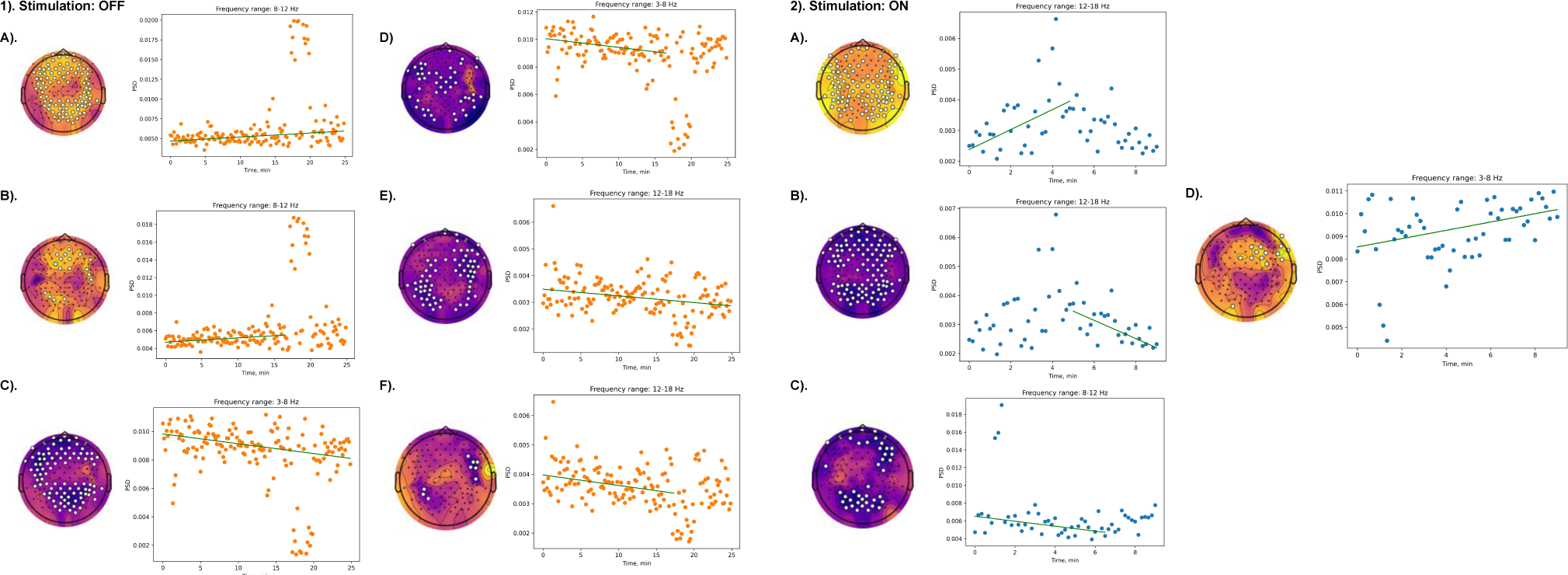
Gradual changes in mean PSD for different frequency ranges in patient P01 induced following PNS offset (1) and onset (2). Green line indicates the mean trend, channels at which significant trends were observed are marked with white dots. Scalp topographies depict Kendalls Tau value

**Fig. 3.**
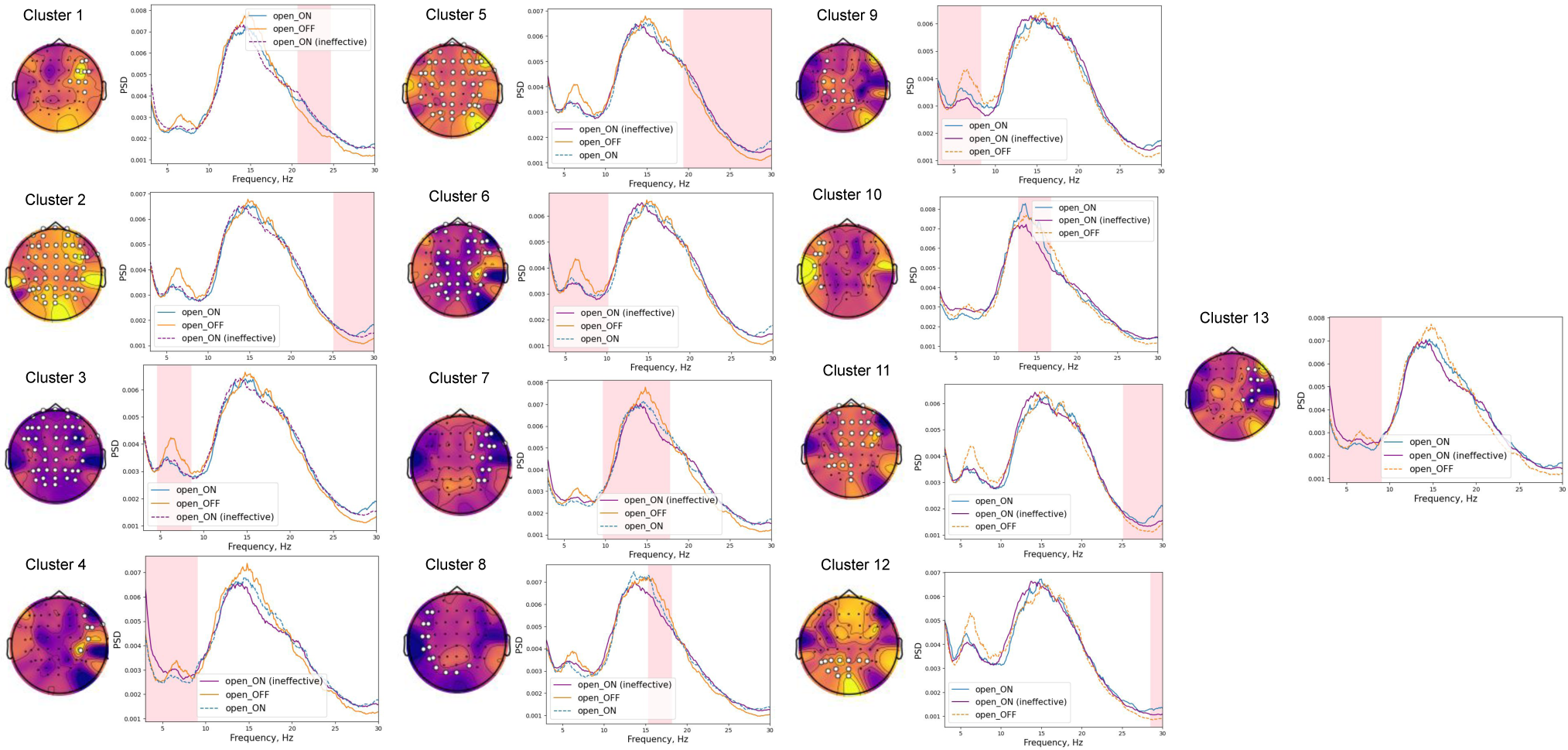
Spatio-spectral clusters in patient P02 with significant differences related to the presence of TENS or PNS. Data for the eyes-open condition are shown. Frequency intervals with significant changes in spectral power are shaded in pink. EEG channels, forming the clusters with significant differences are marked with white dots. Scalp topographies depict the t-values. For visualization purpose, dashed lines represent the data for the conditions that were not used in the given pairwise statistical comparison

The opposite linear trend was observed for theta power (Figure 2, 1.C): it significantly decreased for the whole period of PLP increase at 92 channels (mean slope: -1.151e-05, mean intercept:0.0098). The localization of this decrease corresponded to localization of theta power decrease in stationary conditions (clusters 1 and 2 in Figure 1). The analysis of the period before the sharp decrease of theta power (Figure 2, 1.D) demonstrated that the significant trend was preserved at 47 channels (mean slope: -1.057e-05, intercept: 0.0101).

A linear trend was also found for the low beta power. Mean PSD gradually decreased (mean slope: -4.173e-06, mean intercept: 0.0035) for 67 channels after PNS was turned off (Figure 2, 1.E). This trend was also present after the period of sharp alpha increase was excluded from the analysis (mean slope: -5.944e-05, mean intercept: 0.004). This early trend was significant for 9 channels that corresponded to lateral posterior sites ipsilateral to the amputation and temporal sites contralateral to the amputation (Figure 2, 1.F).

Gradual changes in EEG patterns were also found for the condition where PNS was turned on corresponding to the period during which PLP was fully suppressed. The patient reported that PLP was relieved in 5 min after PNS onset. During this 5-minutes period, a significant gradual increase occurred in low beta power (mean slope: 5.431e-05, mean intercept: 0.0024) as evident from the data of 96 channels primarily localized in posterior scalp regions (Figure 2, 2.A). This significant increase in low beta power was then followed by the decrease (Figure 2, 2.B) with the same spatial properties observed at 105 channels (mean slope: -5.246e-05, mean intercept:0.0035).

A significant decrease in alpha power (mean slope: -4.642e-05, mean intercept: 0.0065) was observed for 42 channels representing the anterior and posterior regions (Figure 2, 2.C). Additionally, a significant increase in theta power (Figure 2, 2.D) was observed at 14 frontal, temporal and central channels contralateral to the side with amputation (mean slope: 3.048e-05, mean intercept: 0.0085).

In patient P02, we analyzed the changes in EEG patterns for both TENS, which was effective to relieve PLP, and PNS, which was ineffective. In this patient, the power spectral density (PSD) exhibited peculiar characteristics, with a prominent high-frequency component ranging from 10 to 30 Hz and a peak at around 6-8 Hz, which could have represented theta activity.

For the eyes-open conditions, the statistical testing revealed 13 significant spatio-spectral clusters (see Figure3 and Table **??**). The comparison of the TENS on and TENS off conditions showed that the 6-8 Hz peak was stronger in the absence of TENS, that is when PLP was present. This effect was evident for the cluster 3, characterized by generalized spatial pattern. The same effect was observed for comparison with the PNS being turned on (cluster 6). At the same time, the increase of aperiodic component in theta range was observed ipsilaterally to the side with amputation, when PNS was turned on (clusters 4 and 13). This effect, as indicated by these two clusters, is specifically associated with ineffective PNS stimulation, as evidenced by its comparison to both PLP- and TENS-related conditions. Simultaneously, this effect was complemented by a reduction in theta power at central sites contralateral to the amputated side (cluster 9). This decrease was observed during ineffective stimulation (PNS) as compared to effective stimulation (TENS).

The activity in higher frequency bands also changed in patient P02 depending on the presence of neurostimulation. Stimulation-related effects were found for high beta power 25-30 Hz, which increased at the anterior and posterior sites as was evident from cluster 1, including fronto-temporal channels ipsilateral to the side of amputation, cluster 2 and 5 with widespread spatial patterns. Additionally, cluster 11 with mostly fronto-central and cluster 12 with mostly posterior localizations demonstrated that effective TENS stimulation induced increase of this high-frequency beta power compared to ineffective PNS stimulation.

The observed effects within the approximately 10-20 Hz frequency range included a decrease in the 9.7 - 17.8 Hz range at channels ipsilateral to the amputated side when the ineffective PNS stimulation was activated, in comparison to the PLP condition (cluster 7). Similarly, decreases in the 15.3 - 18.2 Hz and 12.7 - 16.8 Hz ranges were noted contralaterally when comparing the PNS condition to the PLP condition (cluster 8) and the TENS condition (cluster 10). These differences could be explained not only by power modulations per se, but by the shifts in frequency domain towards the lower frequencies, which were observed only for ineffective PNS stimulation.

For the eyes-closed condition, spectral changes were also found that depended on the presence of PNS or TENS (Fig. 4, Tab. 3). TENS-related increase in aperiodic theta component was observed in clusters 1 and 2, when compared to PLP-related condition. The localization involved frontal and posterior regions. A similar increase in theta activity was observed with PNS, which proved ineffective for pain suppression (clusters 5 and 12). Notably, this PNS-related increase in theta was predominantly bilateral when compared to the TENS condition.

**Fig. 4.**
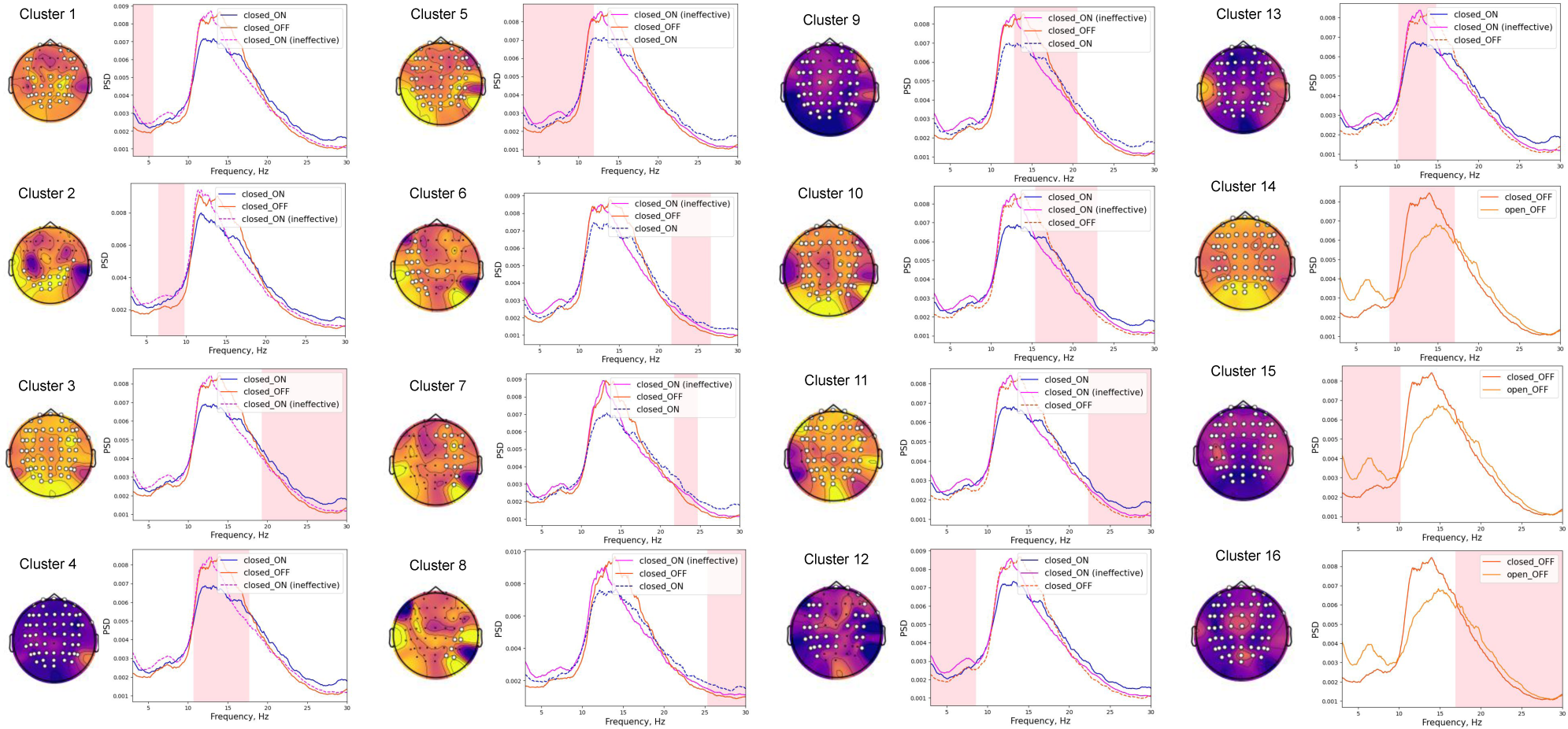
Spatio-spectral clusters in patient P02 with significant differences related to the presence of TENS or PNS. Data for the eyes-closed condition are shown. Frequency intervals with significant changes in spectral power are shaded in pink. EEG channels, forming the clusters with significant differences are marked with white dots. Scalp topographies depict the t-values. For visualization purpose, dashed lines represent the data for the conditions that were not used in the given pairwise statistical comparison

**Table 3.**
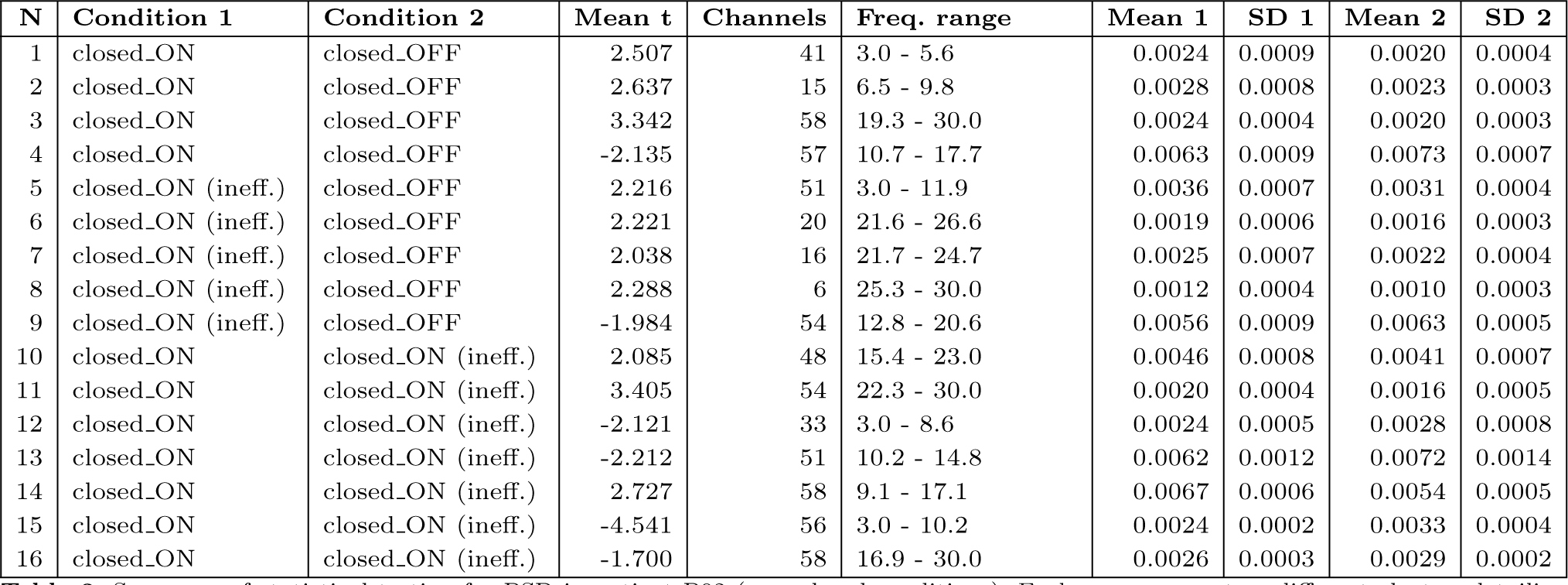
Summary of statistical testing for PSD in patient P02 (eyes-closed conditions). Each row represents a different cluster, detailing the conditions compared (Condition 1 and Condition 2), mean t-values, number of channels involved, frequency range of the cluster, and the mean with standard deviation for both conditions. ‘closed ON‘ condition represents TENS turned on, while ‘closed ON (ineff.)‘ represents PNS, which was ineffective for PLP suppression, turned on

| N | Condition 1 | Condition 2 | Mean t | Channels | Freq. range | Mean 1 | SD 1 | Mean 2 | SD 2 |
| --- | --- | --- | --- | --- | --- | --- | --- | --- | --- |
| 1 | closed_ON | closed_OFF | 2.507 | 41 | 3.0 - 5.6 | 0.0024 | 0.0009 | 0.0020 | 0.0004 |
| 2 | closed_ON | closed_OFF | 2.637 | 15 | 6.5 - 9.8 | 0.0028 | 0.0008 | 0.0023 | 0.0003 |
| 3 | closed_ON | closed_OFF | 3.342 | 58 | 19.3 - 30.0 | 0.0024 | 0.0004 | 0.0020 | 0.0003 |
| 4 | closed_ON | closed_OFF | -2.135 | 57 | 10.7 - 17.7 | 0.0063 | 0.0009 | 0.0073 | 0.0007 |
| 5 | closed_ON (ineff.) | closed_OFF | 2.216 | 51 | 3.0 - 11.9 | 0.0036 | 0.0007 | 0.0031 | 0.0004 |
| 6 | closed_ON (ineff.) | closed_OFF | 2.221 | 20 | 21.6 - 26.6 | 0.0019 | 0.0006 | 0.0016 | 0.0003 |
| 7 | closed_ON (ineff.) | closed_OFF | 2.038 | 16 | 21.7 - 24.7 | 0.0025 | 0.0007 | 0.0022 | 0.0004 |
| 8 | closed_ON (ineff.) | closed_OFF | 2.288 | 6 | 25.3 - 30.0 | 0.0012 | 0.0004 | 0.0010 | 0.0003 |
| 9 | closed_ON (ineff.) | closed_OFF | -1.984 | 54 | 12.8 - 20.6 | 0.0056 | 0.0009 | 0.0063 | 0.0005 |
| 10 | closed_ON | closed_ON (ineff.) | 2.085 | 48 | 15.4 - 23.0 | 0.0046 | 0.0008 | 0.0041 | 0.0007 |
| 11 | closed_ON | closed_ON (ineff.) | 3.405 | 54 | 22.3 - 30.0 | 0.0020 | 0.0004 | 0.0016 | 0.0005 |
| 12 | closed_ON | closed_ON (ineff.) | -2.121 | 33 | 3.0 - 8.6 | 0.0024 | 0.0005 | 0.0028 | 0.0008 |
| 13 | closed_ON | closed_ON (ineff.) | -2.212 | 51 | 10.2 - 14.8 | 0.0062 | 0.0012 | 0.0072 | 0.0014 |
| 14 | closed_ON | closed_ON (ineff.) | 2.727 | 58 | 9.1 - 17.1 | 0.0067 | 0.0006 | 0.0054 | 0.0005 |
| 15 | closed_ON | closed_ON (ineff.) | -4.541 | 56 | 3.0 - 10.2 | 0.0024 | 0.0002 | 0.0033 | 0.0004 |
| 16 | closed_ON | closed_ON (ineff.) | -1.700 | 58 | 16.9 - 30.0 | 0.0026 | 0.0003 | 0.0029 | 0.0002 |

We also found increases in high-frequency components during both TENS and PNS. These effects are represented by clusters 3, 6, 7, 8, 10, and 11, affecting widespread areas. The effect was most pronounced during effective TENS stimulation (clusters 3, 10, and 11). Moreover, ineffective PNS stimulation exhibited a decrease in the 15-20 Hz range compared to both PLP and TENS conditions, revealing a similar shift toward lower frequencies akin to what was observed in eyes-open conditions (clusters 9 and 10).

Finally, TENS decreased power in the 10-15 Hz band compared to both PLP and PNS conditions, as evident from clusters 4 and 13 with widespread spatial patterns.

It should be noted that PSD of patient P02 manifested non-trivial modulations in the 10-20 Hz range. This frequency range is traditionally associated with the beta range, yet the observed modulations were more characteristic of the alpha range. As evident from cluster 14, low-beta power between 9.1 and 17.1 Hz was increased when the eyes were closed eyes-open condition. Additionally, we observed a theta power (3-10.2 Hz) decrease when eyes were closed in cluster 15. Finally, when the eyes were open, the power between 16.9 and 30 Hz was increased as well, resembling the typical behavior of beta rhythm. These observations were taken into account for the subsequent interpretation of the results.

Like in patient P01, the analyses of gradual changes in EEG patterns in patient P02 showed significant trends for the periods following neurostimulation onset or offset (Fig. 5). The observed trends in gradual pain suppression or increase were associated with the modulation of theta power. When the stimulation was turned off and the pain intensified, theta power gradually increased in 30 channels (mean slope 4.071e-05, mean intercept 0.0038) (see Fig. 5 1.A). Conversely, it decreased (mean slope -6.989e-06, intercept 0.0039) mostly at the sites ipsilateral to the amputation (31 channels in total), when stimulation was turned on (Fig. 5 2.A). Notably, during the last several minutes of the recording, this decrease was most pronounced, affecting a total of 34 channels mostly contralateral to the side of amputation (mean slope -3.072e-05, mean intercept 0.0042) (Fig. 5 1.B).

**Fig. 5.**
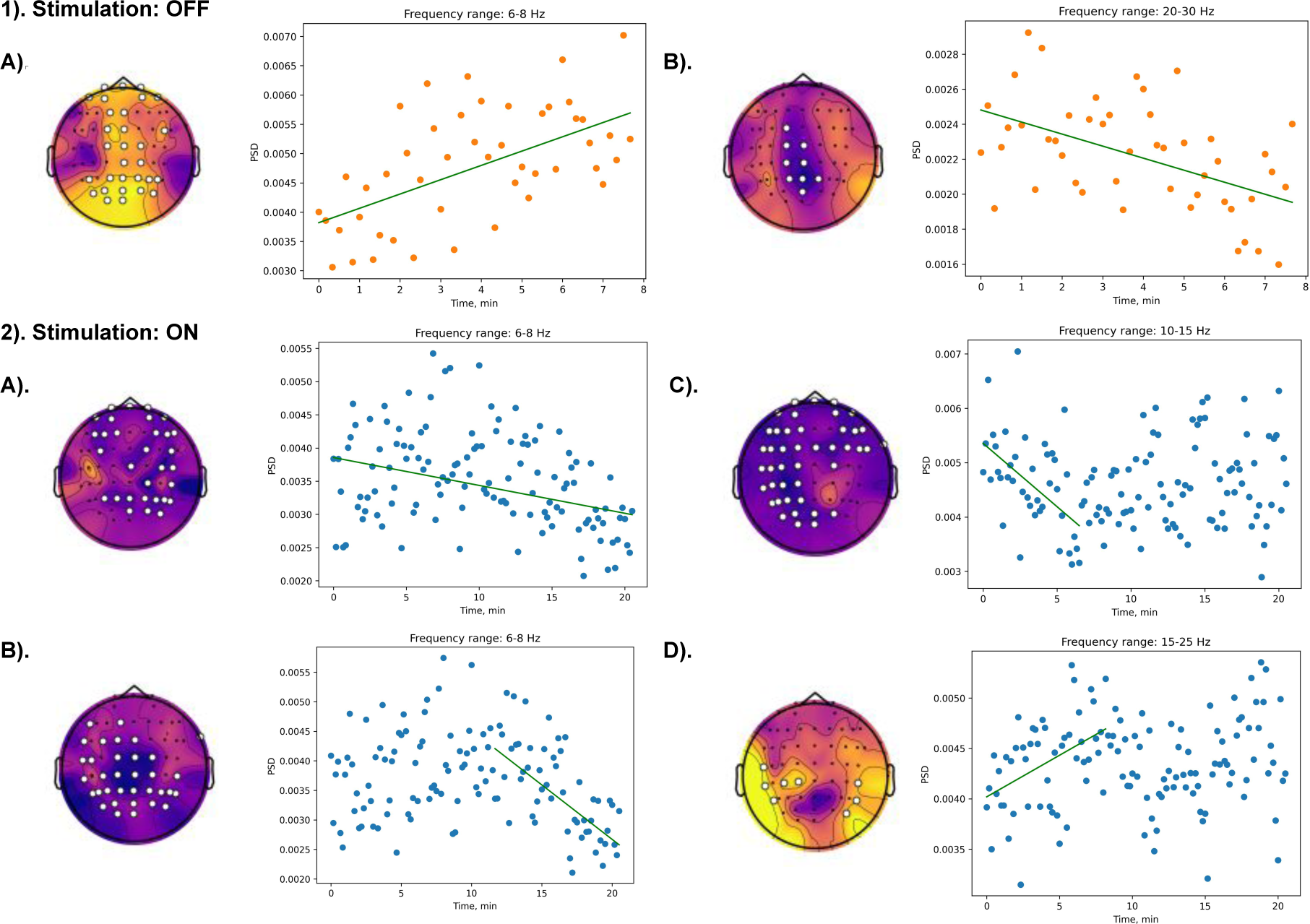
Gradual changes in mean PSD for different frequency ranges in patient P02 during the period when PLP increased after TENS was turned off (1) and decreased after TENS was turned on (2). Green line indicates the mean trend. Channels with significant trends are marked with white dots. Scalp topographies depict Kendalls Tau value.

When the TENS stimulation was turned off and the pain increased, the power in high-frequency range (20-30 Hz) gradually decreased, which was observed at 9 posterior and central channels with the mean slope of -1.146e-05 and mean intercept 0.0025 (see Fig. 5 1.B). Conversely, when the stimulation was turned on and the pain was suppressed, the gradual increase of power between 15 and 25 Hz was observed for the first several minutes at 8 posterior channels, the larger part of which were presented contralateral to the side of amputation (mean slope: 1.376e-05, mean intercept: 0.0040) (see Fig. 5 1.D). In 10-15 Hz range, the short-term stimulation-related decrease was observed at 43 channels mostly contralateral to the side of amputation (mean slope: -3.880e-05, intercept: 0.0054) (see Fig. 5 1.C).

In patient P03 (Fig. 6 and Tab. 4 ) (who for some period of time experienced pain suppression without stimulation due to analgetics), EEG patterns changed in association with SCS being turned on or off or irrespective of SCS.

**Fig. 6.**
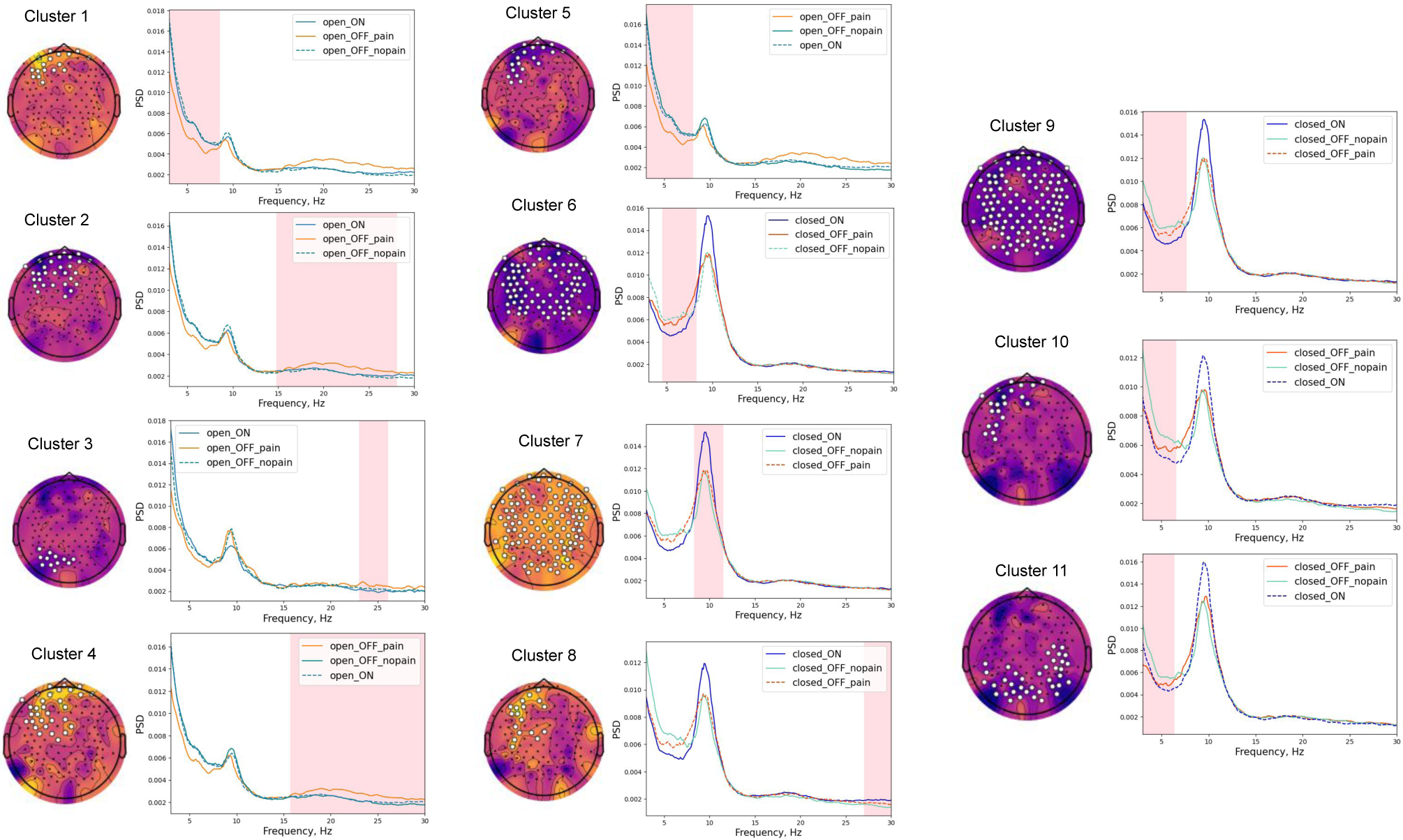
Spatio-frequency clusters with significant changes associated with the presence or absence of SCS in patient P03. Frequency intervals with significant changes in spectral power are shaded in pink. EEG channels forming the clusters with significant differences are marked with white dots. Scalp topographies depict t-values. For comparison purposed, the dashed line represents the conditions that was not used in the particular statistical analyses

**Table 4.** Summary of statistical testing for PSD in patient P03. Each row represents a different cluster, detailing the conditions compared (Condition 1 and Condition 2), mean t-values, number of channels involved, frequency range of the cluster, and the mean with standard deviation for both conditions.

| N | Condition 1 | Condition 2 | Mean t | Chan. | Freq. range | Mean 1 | SD 1 | Mean 2 | SD 2 |
| --- | --- | --- | --- | --- | --- | --- | --- | --- | --- |
| 1 | open_ON | open_OFF (pain) | 2.324 | 17 | 3.0 - 8.6 | 0.0076 | 0.0016 | 0.0061 | 0.0015 |
| 2 | open_ON | open_OFF (pain) | -1.934 | 30 | 14.8 - 28.1 | 0.0023 | 0.0005 | 0.0028 | 0.0006 |
| 3 | open_ON | open_OFF (pain) | -2.665 | 12 | 23.0 - 26.1 | 0.0020 | 0.0006 | 0.0026 | 0.0006 |
| 4 | open_OFF (pain) | open_OFF (no pain) | 2.362 | 31 | 15.7 - 30.0 | 0.0027 | 0.0006 | 0.0022 | 0.0004 |
| 5 | open_OFF (pain) | open_OFF (no pain) | -2.649 | 20 | 3.0 - 8.2 | 0.0063 | 0.0015 | 0.0081 | 0.0010 |
| 6 | closed_ON | closed_OFF (pain) | -2.480 | 80 | 4.5 - 8.3 | 0.0053 | 0.0010 | 0.0064 | 0.0013 |
| 7 | closed_ON | closed_OFF (no pain) | 2.346 | 106 | 8.3 - 11.5 | 0.0110 | 0.0033 | 0.0088 | 0.0028 |
| 8 | closed_ON | closed_OFF (no pain) | 2.806 | 18 | 27.0 - 30.0 | 0.0019 | 0.0007 | 0.0015 | 0.0003 |
| 9 | closed_ON | closed_OFF (no pain) | -2.858 | 115 | 3.0 - 7.7 | 0.0055 | 0.0012 | 0.0068 | 0.0013 |
| 10 | closed_OFF (pain) | closed_OFF (no pain) | -2.409 | 21 | 3.0 - 6.6 | 0.0064 | 0.0016 | 0.0079 | 0.0014 |
| 11 | closed_OFF (pain) | closed_OFF (no pain) | -2.209 | 38 | 3.0 - 6.4 | 0.0054 | 0.0018 | 0.0067 | 0.0017 |

For the eyes-open condition, the pain-related effects were observed at fronto-central sites ispilateral to the amputation (clusters 1 and 5) with PSD decrease observed for the the theta range when SCS was turned off and the patient felt PLP. Additionally, decrease of aperiodic theta component, when the pain was present compared to non-painful stimulation-free condition, was observed in frontal channels ipsilateral to the stimulation side (cluster 10) and posterior channels (cluster 11).

For cluster 4 that demonstrates the comparison of pain presence and pain absence, when the stimulation was turned off, PSD decreased in broad beta range from 15.7 to 30 Hz at ipsilateral frontal sites, when the pain was present. Additionally, in comparison to stimulation turned on, a pain-related increase in broad beta power was observed for cluster 2 and for cluster 3 involving frontal and posterior regions ipsilateral to the side of amputation.

For the eyes-closed condition, SCS modulated EEG patterns even without modulating PLP. For clusters 6 and 9 characterized by widespread spatial patterns mean PSD of aperiodic theta component decreased, when the SCS was turned on in comparison to both painful and non-painful stimulation-free conditions. Additionally, alpha power increased in a generalized manner when SCS was turned on compared to the absence of pain, when the stimulation was turned off (cluster 7). For the comparison of the same conditions, the power of 27-30 Hz range increased in frontal channels ipsilateral to the amputation side (cluster 8).

### SSD patterns

To investigate the potential sources of the identified spatio-frequency components, we conducted a spectro-spatial decomposition of periodic parts of PSD (Fig. 7). This analysis showed that the theta-frequency components observed in participants P01 and P02 (Fig. 7 1.A and 2.A) and alpha activity in patient P03 (Fig. 7 3.A). originated from the parieto-occipital regions. Furthermore, low beta activity in patient P01, which increased during PNS, was localized in the occipital regions contralateral to the amputation (Fig. 7 1.C). Yet, pain-related beta activity in patient P03 was localized at the fronto-temporo-central regions ipsilateral to the amputation (Fig. 7 3.B). Notably, in patient P01, the localization of alpha activity was contralateral to the amputation in the presence of PLP (Fig. 7 1.A).

**Fig. 7.**
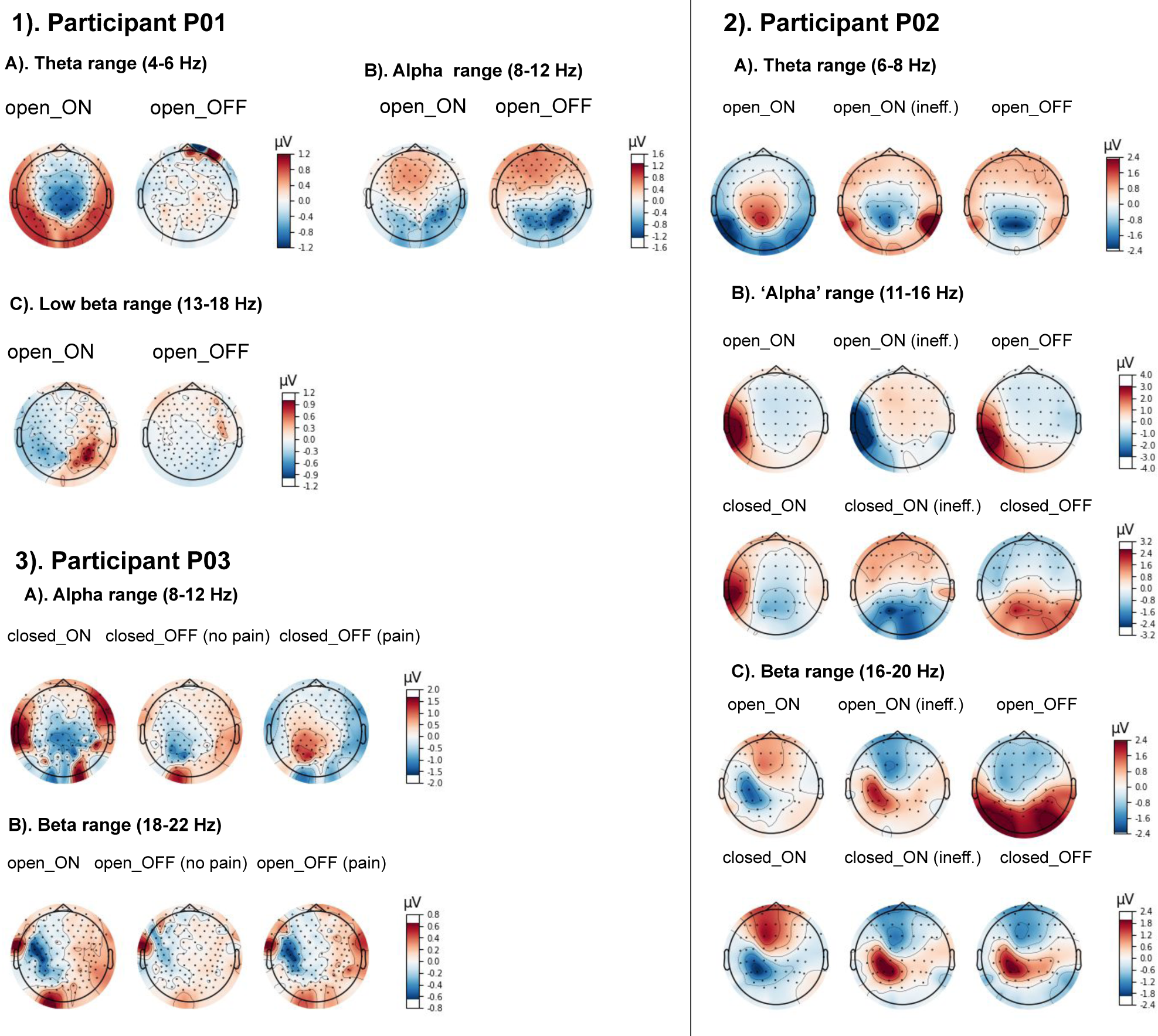
Spatial patterns in frequency bands for each participant and condition

Additionally, patient P02 had a distinctive pattern where presumably alpha oscillations (11-16 Hz) were localized in temporal regions contralateral to the amputation, whereas in eyes-closed conditions these oscillations were predominantly located in parieto-occipital regions, except the case when TENS was turned on (Fig. 7 2.B). Beta-range (16-20 Hz) oscillations exhibited a somatosensory topography contralateral to the amputated side (see Fig. 7 2.C). This pattern persisted across all conditions, with the exception of when the stimulation was turned off and the eyes were open. In this specific case, the topography indicated the involvement of posterior regions.

### Fractal dimension

Fractal dimension (FD) analysis assessed EEG signal complexity (Fig. 8 and Tab. 5).

**Fig. 8.**
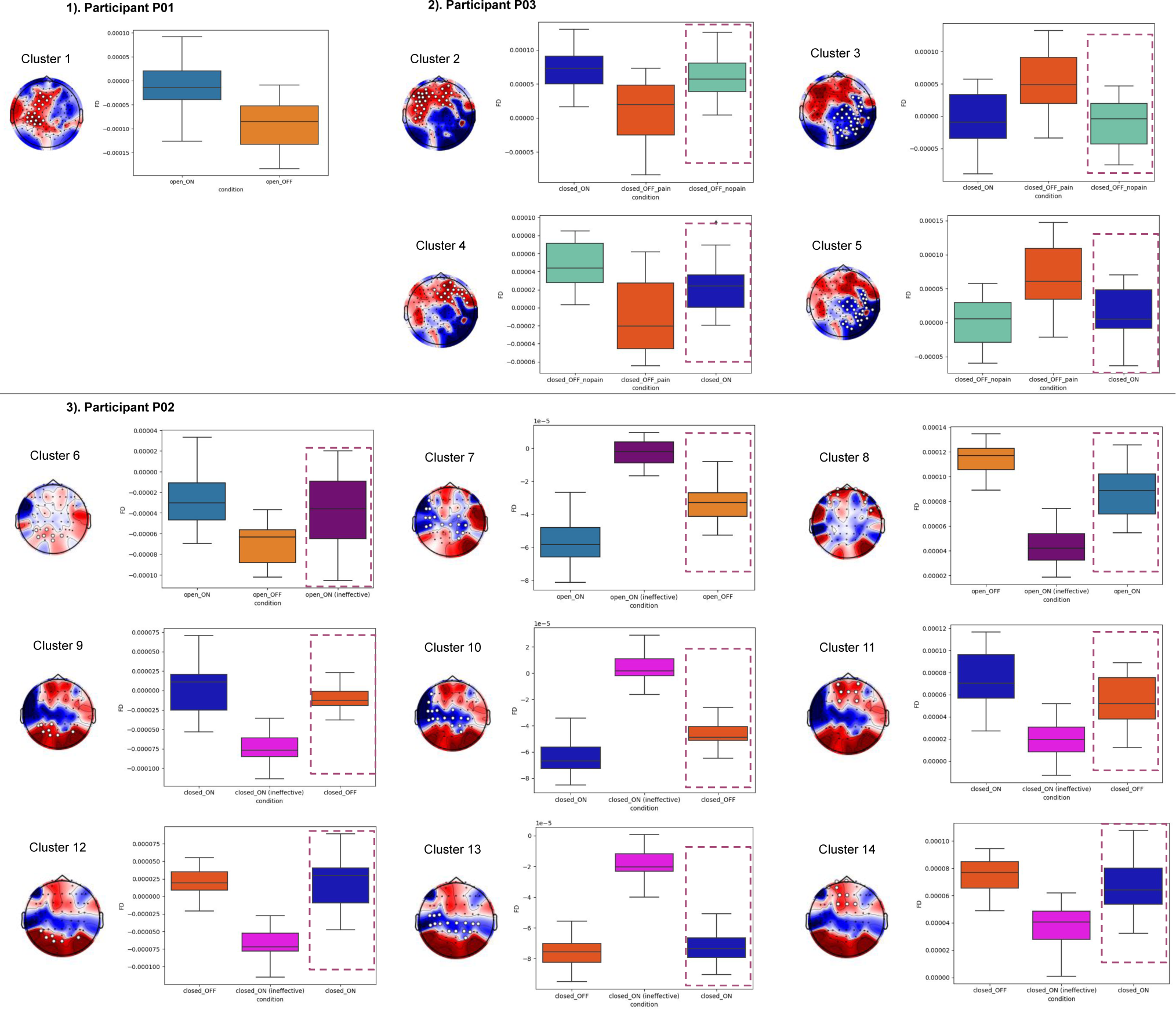
The analysis of fractal dimensions. Clusters with significant across-condition differences are shown. EEG channels, forming the clusters with significant differences, are marked with white dots. Scalp topographies depict t-values. The dashed rectangles represent the conditions which were not entered in the statistical analysis

**Table 5.** Summary of statistical testing for FD in all three patients. Each row represents a different cluster, detailing the patient, conditionscompared (Condition 1 and Condition 2), mean t-values, number of channels involved, and the mean with standard deviation for both conditions.

| N | Patient | Condition 1 | Condition 2 | Mean t-value | Chan. | Mean 1 | SD 1 | Mean 2 | SD 2 |
| --- | --- | --- | --- | --- | --- | --- | --- | --- | --- |
| 1 | P01 | open_ON | open_OFF | 2.961 | 19 | -0.000013 | 0.000046 | -0.000096 | 0.000050 |
| 2 | P03 | closed_ON | closed_OFF (pain) | 3.580 | 29 | 0.000074 | 0.000030 | 0.000007 | 0.000048 |
| 3 | P03 | closed_ON | closed_OFF (pain) | -3.518 | 31 | -0.000007 | 0.000041 | 0.000056 | 0.000044 |
| 4 | P03 | closed_OFF (no pain) | closed_OFF (pain) | 3.979 | 21 | 0.000047 | 0.000026 | -0.000010 | 0.000042 |
| 5 | P03 | closed_OFF (no pain) | closed_OFF (pain) | -4.720 | 30 | 0.000001 | 0.000034 | 0.000072 | 0.000045 |
| 6 | P02 | open_ON | open_OFF | 4.153 | 7 | -0.000026 | 0.000028 | -0.000068 | 0.000018 |
| 7 | P02 | open_ON | open_ON (ineff.) | -5.529 | 16 | -0.000056 | 0.000013 | -0.000003 | 0.000008 |
| 8 | P02 | open_ON | open_ON (ineff.) | 9.271 | 10 | 0.000115 | 0.000012 | 0.000043 | 0.000014 |
| 9 | P02 | closed_ON | closed_ON (ineff.) | 7.584 | 8 | 0.000003 | 0.000032 | -0.000075 | 0.000019 |
| 10 | P02 | closed_ON | closed_ON (ineff.) | -7.266 | 16 | -0.000065 | 0.000012 | 0.000004 | 0.000010 |
| 11 | P02 | closed_ON | closed_ON (ineff.) | 6.219 | 7 | 0.000073 | 0.000025 | 0.000020 | 0.000015 |
| 12 | P02 | closed_OFF | closed_ON (ineff.) | 9.611 | 6 | 0.000022 | 0.000019 | -0.000069 | 0.000022 |
| 13 | P02 | closed_OFF | closed_ON (ineff.) | -6.170 | 16 | -0.000076 | 0.000010 | -0.000018 | 0.000010 |
| 14 | P02 | closed_OFF | closed_ON (ineff.) | 4.640 | 9 | 0.000074 | 0.000012 | 0.000037 | 0.000016 |

In patients P01 and P03, effects of PLP suppression was associated with an increase of FD at the sites ipsilateral to the amputation side (clusters 1, 2 and 4). Additionally, the decrease of FD at the contralateral channels were observed in patient P03 with respect to PLP suppression.

In patient P02, effective TENS stimulation induced increase of FD at posterior channels in eyes-open condition (cluster 6). The ineffective PNS stimulation induced decrease of FD at the same regions compared to effective TENS stimulation (cluster 9) and PLP presence (cluster 12). Additional decrease of FD for ineffective PNS stimulation was observed at fronto-central sites (clusters 8, 11, and 14). Similarly, in line with the trends seen in patients P01 and P03, there was a decrease in FD at channels mostly contralateral to the amputated side when comparing effective TENS to ineffective PNS. Moreover, the FD was higher in the ineffective TENS condition than in the PLP presence condition under eyes-closed circumstances, involving bilateral temporal regions and centro-parietal regions (cluster 13).

## Discussion

In this study, we investigated the effects of PLP-relieving neurostimulation on the EEG patterns in three amputees. Resting-state EEG was recorded in several conditions, including PLP presence in the absence on neurostimulation, its absence in the presence of neurostimulation and a case of neurostimulation which was not effective to relieve PLP (PNS in P02). Overall, distinct EEG patterns were found for each condition, with a number of individual differences. EEG of each patient exhibited unique characteristics in terms of power spectrum density (PSD) and its modulations related to the presence of neurostimulation and its efect on PLP. Having the eyes open or closed also contributed to the individual variability.

The neurostimulation/PLP-sensitive EEG components were widespread over the cortical surface rather than corresponding to focal somatosensory or motor representations. Despite the across-participant variability, several common effects were observed, which were consistent with the previous literature. Thus, beta-band activity was elevated in the frontal regions ipsilateral to the amputation in two patients (P01 and P03) during the presence of PLP. In patient P01 this elevation was observed in the range 25-30 Hz and in patient P03 in the range 18-30 Hz. These findings match the previous reports of increased high-beta activity in different pain syndromes (Vanneste et al., 2017; Wang et al., 2019). Such an increased beta activity could reflect an alteration in the descending pain inhibition pathway linked to lateral prefrontal regions (Bräscher et al., 2016).

Another effect similar in patients P01 and P03 is decrease of aperiodic theta component, when the pain is present. It is supposed that changes in aperiodic brain activity can indicate shifts in the balance of excitatory and inhibitory neural activities (Gao et al., 2017). Specifically, a steeper spectral exponent is believed to signify a greater influence of inhibitory over excitatory activity, whereas a shallower spectrum indicates the opposite. Increases in excitatory activity are associated with decreased synchronization between oscillatory cycles and neuron spikes, leading to higher neural noise that potentially disrupts neural communication. Thus, elevations in broadband power are related to increased rates of neuron firing. This increase may contribute to the decoupling of neuronal spikes from neural oscillations, thereby introducing additional noise in the neural system (Voytek and Knight, 2015). The observed pain-related reduction of aperiodic theta activity in frontal cortex is in line with previously reported reduction in excitatory inputs from the thalamus to the medial prefrontal cortex (mPFC) in chronic pain models, which affects both pyramidal neurons and inhibitory interneurons (Jefferson et al., 2021). Furthermore, this finding is consistent with the increase in frontal theta band activity observed following chronic pain relief, as reported by (Rustamov et al., 2022).

In patient P03 the mentioned increase in beta power and decrease in theta are purely pain-related effects, as they were not observed both in stimulation-free non-painful conditions and while SCS was turned on.

In each patient, we observed distinct and specific effects manifesting the neurostimulation and PLP conditions. Thus, increases in alpha power were prominent in patient P01 during the presence of PLP. PLP in patient P01 was accompanied with a feeling of stiffness, which bears relevance to the report of Simis et al. (2022a), where alpha band activity and stiffness were positively correlated in patients with osteoarthritis, which could reflect the influence of cortical inhibition due to peripheral damage. Furthermore, studies of peripheral neurogenic pain revealed modulations of the resting-state alpha or theta activity (Sarnthein et al., 2006; Olesen et al., 2011; van den Broeke et al., 2013). Similar EEG changes were observed for non-neuropathic painful conditions (Case et al., 2017; Fallon et al., 2018; Dinh et al., 2019), although some of them were related to pain chronification rather than pain intensity per se.

Thalamocortical dysrhythmia (TCD) is one explanation for the alterations in theta and alpha rhytms in nociception (Jeanmonod et al., 2001). According to this framework, transfer of thalamic bursts of activity to the cortex acts as an abnormal painful input. This framework usually considers theta oscillations alone. However, since the alpha rhythm is a part of activity in thalamocortical loops, this explanation could be extended to the alpha range, as well. In this respect, the absence of typical alpha activity and prominent 10-15 Hz oscillations observed in patient P02 could also represent thalamocortical oscillations altered during the presence of PLP. An increase of the power of this during the eyes closed condition in this patient suggests that this activity could have been a shifted alpha rhythm. On the other hand, this could have also been the result of the edge effect (Llińas et al., 1999), where high-frequency oscillations are generated because of a pronounced pathological low-frequency activity. In patient P02 this low-frequency activity could have been represented by a pronounced theta peak. Similar increase in the theta and beta oscillations have been reported in patients with chronic pain (Zebhauser et al., 2023). Thus, in patient P02, TENS could have suppressed an abnormal low-frequency thalamocortical activity.

On the other hand, it should be noted that the standard theta-component observed in eyes-open condition in patient P02, was increased when the stimulation was turned off and suppressed irrespective of the type of stimulation (effective TENS and ineffective PNS). The observation of this pronounced pain-related theta peak is not in conflict with our previous observations of pain-related decreased aperiodic theta activity in patients P01 and P02, since not all the patients with chronic pain develop abnormal periodic theta oscillations (Jensen et al., 2013; Schmidt et al., 2012; Rustamov et al., 2022). Therefore, the suppression of a pronounced theta peak induced by stimulation and the increase in aperiodic theta activity should be viewed as distinct processes. The former indicates modulations in TCD, while the latter reflects alterations in the balance between excitation and inhibition. The independence of these processes is supported by the fact that aperiodic theta changes in patients P01 and P03 were mainly localized at frontal channels, while pain-related periodic theta component in patient P02 covered widespread regions.

Interestingly, changes in aperiodic theta activity induced by stimulation were also observed in Patient P02. This included ipsilateral fronto-temporal channels in the eyes-open condition and lateralized wide regions in the eyes-closed condition, but only for ineffective PNS. Effective TENS led to an increase in aperiodic theta activity, primarily localized in posterior regions. This pattern may indicate general alterations in excitation specific to the stimulation itself, rather than effects related to pain relief.

Changes in alpha activity, which is usually the most prominent EEG oscillation, were also observed. In patient P01, alpha power decreased during PNS, and this decrease was accompanied by an increase in low-beta activity (12-17 Hz). The latter effect is consistent with the previous literature that reported suppression of beta oscillations, reflecting idle state of sensory and motor systems, in pain conditions (Simis et al., 2022b; Teixeira et al., 2021). Additionally, neurofeedback aimed at increasing the low-beta oscillations is effective to treat chronic neuropathic pain (Hassan et al., 2015; Vučkovíc et al., 2019). The mechanisms underlying the interrelation between beta rhythm and nociception could include GABAergic signaling (Barr et al., 2013). In this interpretation, pain is caused by an imbalance between excitation and inhibition with a bias towards excitation, resulting in a reduction of inhibitory input mediated by GABAergic neursons. Since the mechanisms of beta and gamma oscillations essentially depend on the inhibitory interneurons (Baumgarten et al., 2016), beta power could be considered as an indicator of GABAergic activity. Following this argumentation, the increase in beta power observed in our study could indicate facilitation of GABAergic activation by neurostimulation.

An alpha peak was absent in patient P02 spectra, so changes in pure alpha activity are unclear in this peculiar case, although we suggest them to be related to previously reported alterations in 10-15 Hz band. Patient P03 had a spectral peak at the alpha frequency. Although the effects of SCS in this patient were confounded with the effects of medication, changes in alpha power did occur after SCS was turned on. In the eyes-closed condition, this was an increase in alpha power accompanied by a decrease in theta power. This result is consistent with the previously assessed resting-state spectral characteristics during the application of SCS (Witjes et al., 2023), where several studies reported increase of alpha power. In addition to the possible effect of the medication, differences in the alpha-power patterns in patients P01 and P03, could have been related to stimulation type (PNS versus SCS) and the specifics of stimulation patterns. Burst-stimulation in patient P01 could have had an excitatory effect coupled to a decrease in alpha oscillations, while tonic stimulation in patient P03 could have inhibitory effect resulting in an increase in alpha power (Goudman et al., 2020).

Our interpretation that changes in EEG spatio-spectral characteristics reflected the level of PLP is supported by the observation of gradual changes in the EEG spectral characteristics after neurostimulation was turned off or on and PLP changed gradually. In patient P01, alpha power was increasing and theta and low beta power was decreasing after PNS was turned off and PLP was intensifying. Conversely, after PNS was turned on, PLP was decreasing in patient P01 and alpha power was decreasing and theta and beta power were increasing. Gradual changes in EEG patterns were observed in patient P02, as well. Theta power was increasing and high-beta power was decreasing as PLP was intensifying after TENS was turned off, and theta power with 10-15 Hz oscillations were decreasing and 15-25 Hz band power was increasing after TENS was turned back on, which relieved PLP. This capacity of our approach to not only distinguish between the presence and absence of pain based on EEG signals but also estimate the gradual changes in PLP is a valuable outcome for the development of prediction systems within the context of closed-loop neurostimulation systems.

The patient-specific variations in the EEG spectral characteristics highlight the need for a comprehensive understanding of each patient’s spectral profile. They also present a challenge in obtaining universal markers of PLP and its suppression by neurostimulation. Yet, FD, the metric of signal complexity showed more consistency for all patients. An increase in FD on the side contralateral to the amputation or decrease of FD on the side ipsilateral to the amputation was associated with the presence of

PLP, while a decrease in FD on the contralateral side or increase of FD on the ipsilateral sie was associated with PLP suppression by an effective neurostimulation. This suggests that FD could potentially serve as a valuable and standardized metric for assessing pain and evaluating the impact of neurostimulation. There is no explicit indication of whether absolute increase or decrease in complexity measures corresponds to normalization of CNS activity. It was proposed that the deviation from an ’optimal variability’ could become a criterion for the presence of a disorder (Lau et al., 2022), and the estimation of ranges of complexity measures of brain activity in healthy people and patients with neuropathic pain could form a distinct direction of research.

The main limitation of the study is that the number of participants and heterogeneity of their symptoms and response to the treatment does not allow for general conclusions. A larger sample of participants with more similar symptoms and therapeutic strategies would increase the reliability of the results. The other restrictions of the current design include possible overlap between EEG spectral signatures of conscious sensory experiences resulting from neurostimulation and PLP. This aspect should be controlled with sham stimulation at the same location but another parameters which do not induce pain relief. In our study we analyzed the paradigm with an ineffective stimulation, as it followed the therapeutic search for the effective treatment and did not include this scenario as a control condition for each of the patient.

The other methodological limitation is that the naturalistic experiments with neurostimulation and PLP suppression are associated with the recording of resting-state EEG data, which is not stable by its nature and is not as robust as evoked responses with well-known components calculated from the averaged trials. Indeed, it would be possible to evaluate context-dependent evoked responses to repetitive stimulation, but this paradigm creates the risks for sensory habituation and the observed changes of evoked responses may not reflect the changes in pain intensity (although it is possible to add sham stimulation as a control for this case, as well).

The other limitation is that stimulation-induced sensations could inadvertently compete with the intended pain suppression mechanisms. Addressing this limitation demands a comprehensive approach that considers not only pain reduction but also the broader neural consequences of neurostimulation.

The final limitation of the study is that multimodal nature of pain implies the involvement of multiple cortical regions, and the given the experimental design does not directly address or modulate the activity of the possible regions of interest (e. g. phantom limb representation in sensorimotor cortex, integrative nodes of nociception in insula, operculum, etc.). The use of high-density EEG partly solves this problem, as its high spatial resolution allows for implementing signal decomposition techniques and analyze data-driven regions of interest. Yet, additional preliminary mapping of the areas corresponding to phantom limb representation in the brain of given patient could be beneficial.

Despite the mentioned limitations, our results are generally in line with the previous studies, particularly regarding the consideration of the lower-frequency oscillations (theta and alpha) as markers of PLP and of the beta oscillations as a marker of pain-relieving neurostimulation. Taken together, these findings will likely contribute to the development of closed-loop systems for PLP suppression.

## Data Availability

All data produced in the present study are available upon reasonable request to the authors

## Acknowledgements

This work was supported by the Russian Science Foundation under grant № 21-75-30024.

